# Non-Invasive Systemic Blood Pressure Measurement Wait Period Error Mitigation

**DOI:** 10.1101/2024.10.03.24314772

**Authors:** Seán McMahon, Paul Morrow

## Abstract

World Health Organisation blood pressure measurement guidelines stipulate that a patient should be seated, back straight and legs uncrossed, for five minutes prior to blood pressure measurement. A study shows that in the majority of cases, the wait period is disregarded by the physician and consequently, the patient may be exposed to the risk of missed diagnosis or misdiagnosis. Modern oscillometric blood pressure measurement devices do not enforce a wait period or monitor patients to gauge their readiness for a measurement. This study investigates whether the electrocardiogram (ECG) or photoplethysmography (PPG) data of a patient could be used as a determinant of readiness for blood pressure measurement. The results indicate that the standard deviation of the interpeak difference of a patient’s ECG bio-signal could be used as a trigger for blood pressure measurement. In this single participant study, the wait period could be reduced from five minutes to 35 seconds. A follow up clinical investigation is recommended to validate the result on broader populations and a list of improvements are provided for such a follow up investigation.

## Introduction

The World Heart Organization, (WHO), refers to hypertension as the ‘silent killer’ and describes hypertension as the greatest risk factor for death globally (World Health Organization, 2023). It notes that hypertensive heart disease has risen from the 18th leading cause of death to the 9th leading cause of death in high income countries (World Health Organisation, 2020). However, hypertension is also the leading adjustable risk factor for premature death globally (Mills, et al., 2020) and therefore, the importance of correctly diagnosing disease should not be underestimated.

Diagnosis of hypertension, or many other blood pressure, (BP), related diseases, is achieved by measuring systemic blood pressure. Systemic blood pressure is a measure of the force exerted by blood as it flows through the vascular system due to the pumping action of the heart (Magder, 2018). As distinct from pulmonary BP associated with the lungs, systemic BP occurs when the left ventricle supplies oxygenated blood to systemic circulation and the right atrium receives deoxygenated blood from systemic circulation. As regards hypertension, systolic BP is regarded as normal if it is below 120 mmHg (Fogoros, 2023)..

The field of systemic blood pressure measurement has been well established since Korotkoff identified particular sound patterns during the decompression of an artery from a compressed state (Shevchenko & Tsitlik, 1996).and is now largely dominated by cuff-based BP measurement devices. The WHO has issued technical requirements for cuff-based BP measurement (John, et al., 2021) but do not yet recommend the use of devices based on photoplethysmography or other non-invasive cuff-less solutions.

Despite the serious nature of BP related diseases, the need to diagnose correctly and the established nature of the technology, modern BP measurement devices have significant sources of error associated with them such as psychological, (e.g. White Coat Hypertension), device related (e.g. misfitting cuff) and procedural (e.g. observer error) (Mukkamala, et al., 2022). A PRISMA based meta-analysis of academic papers from Pubmed and IEEE suggests a primary academic focus on cuffless based BP devices and the most common error source being device related.

The current BP measurement recommendation is to remain seated, with back straight for five minutes prior to a BP measurement(James & Gerber, 2018). However, studies indicate individuals with BP levels less than 140mmHg could be measured without a wait period and that implementing such an approach might increase screening volume by approximately 60% (Brady, et al., 2021). This creates a conundrum as BP must be first measured, to determine if it is less than 140mmHg. In fact, a recent study showed in the majority of cases, physicians had abandoned guidelines and were taking immediate measurements potentially putting patients at the risk of misdiagnosis and mistreatment (Levy, et al., 2016).

The objective of this study is to determine if BP measurement error can be mitigated through the dynamic monitoring of patients connected to a BP measurement device so that the optimum time to initiate BP measurement is utilised. A correctly functioning algorithm, should reduce the wait period, especially for those with BP under 140mmHg, and encourage physicians to follow good clinical practice thereby reducing the likelihood of misdiagnosis.

## Materials and Methods

To conduct the primary research, a critical literature review of BP measurement was first carried out including a meta-analysis using PRISMA guidelines of PubMed and IEEE articles whereby the results were reviewed and discussed. Following the confirmatory literature review it was elected to use photoplethysmography (PPG) and electrocardiogram (ECG) technologies to investigate whether an optimum measurement state could be discovered.

### Equipment

Clinical care focussed corporations such as Baxter, Medtronic, General Electric and Omron manufacturing vital signs monitoring devices may require costly electronic healthcare record systems to access the raw signal data, (Hillrom, 2023). Consumer devices from Garmin, Fitbit, etc. are not sold as research devices and it is not clear that it is possible to derive the raw data required to answer the research question. Several electronic component manufacturers have development kits, that enable direct measurement of PPG and ECG human vital signs. Selected kits were scored and the MAX86150EVSYS chosen as per figure 1.

**Figure 1:**
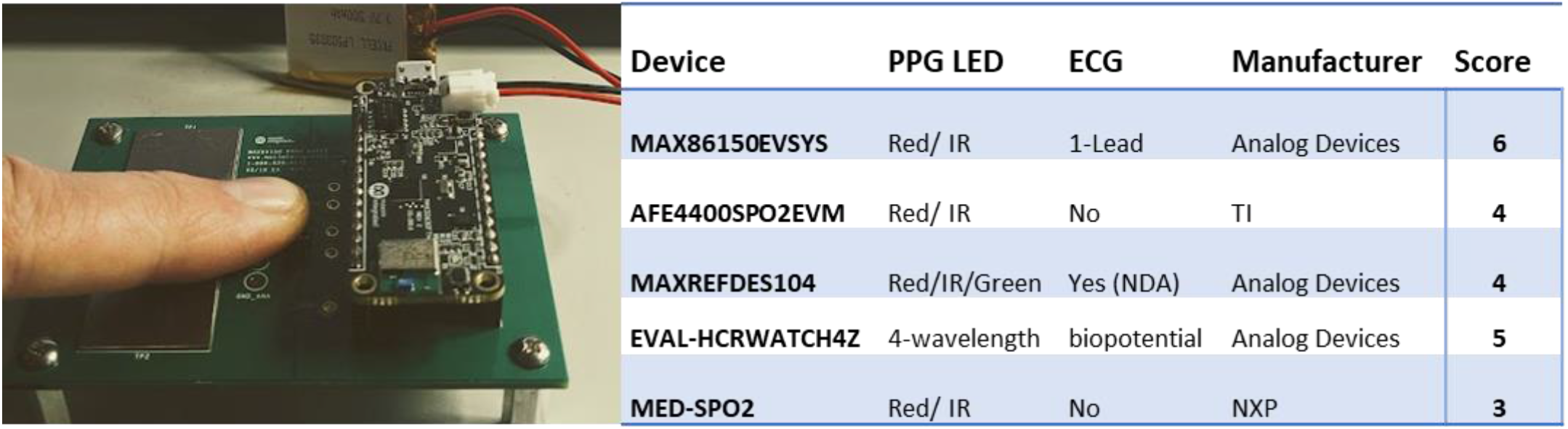
MAX86150EVSYS kit (left) and device selection scoring table (right)

The Omron M3 Comfort, (Model: HEM-7155-E), upper arm BP measurement device was chosen to take BP readings and a Rigel Uni-Sim requiring calibration was used to validate the MAX86150EVSYS ECG function. *Study Protocol*

The single participant clinical investigation study protocol was designed to take 16 separate readings as per table 1, where the second eight reading are repeated with an exercise intervention. The protocol was preregistered on aspredicted.org at https://aspredicted.org/zg4um.pdf and passed ethics approval with the Technological University of the Shannon.

**Table 1:**
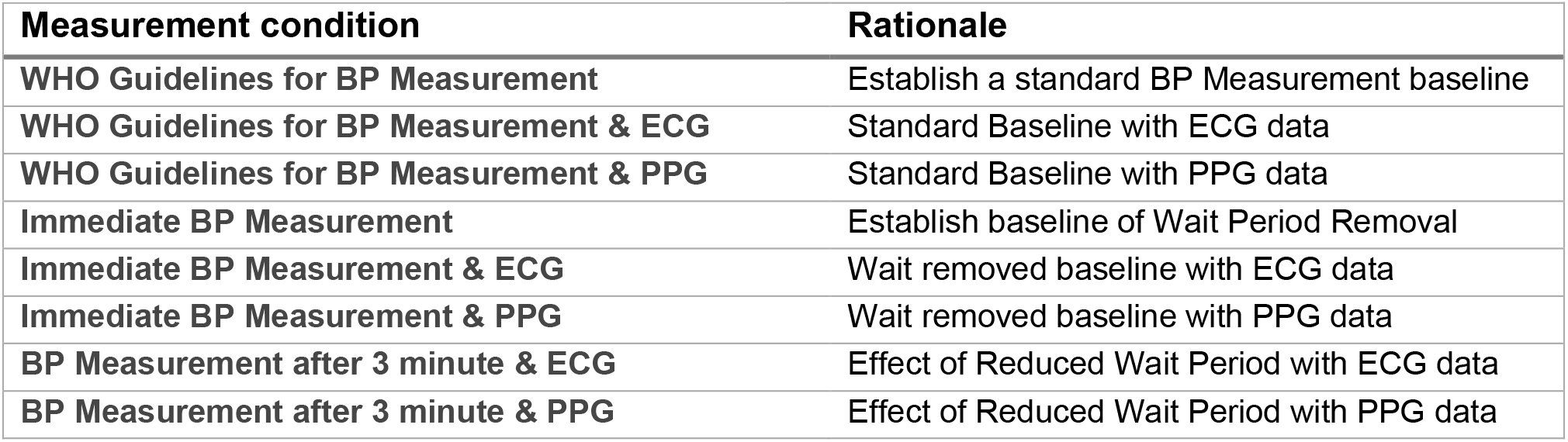
Measurement conditions and rationale.

| Measurement condition | Rationale |
| --- | --- |
| WHO Guidelines for BP Measurement | Establish a standard BP Measurement baseline |
| WHO Guidelines for BP Measurement & ECG | Standard Baseline with ECG data |
| WHO Guidelines for BP Measurement & PPG | Standard Baseline with PPG data |
| Immediate BP Measurement | Establish baseline of Wait Period Removal |
| Immediate BP Measurement & ECG | Wait removed baseline with ECG data |
| Immediate BP Measurement & PPG | Wait removed baseline with PPG data |
| BP Measurement after 3 minute & ECG | Effect of Reduced Wait Period with ECG data |
| BP Measurement after 3 minute & PPG | Effect of Reduced Wait Period with PPG data |

### Code

The study analysis software code was designed to select a window of 4000 ECG or PPG data samples equivalent to 20 seconds of data. The scipy library function signal.find_peaks was called to discover the ECG and PPG Peaks. The number of samples between peaks was computed and saved to an array. The standard deviation (SD) of the array was calculated with the intent of finding an array with a SD less than 9. The window was set to shift by 2.5 seconds as the algorithm searched the data for an array of interpeak intervals that met the criterion. Once the trigger conditions were calculated, as per equation (1), and satisfied, the settled ECG or PPG dataset was said to represent an optimum time for BP measurement.

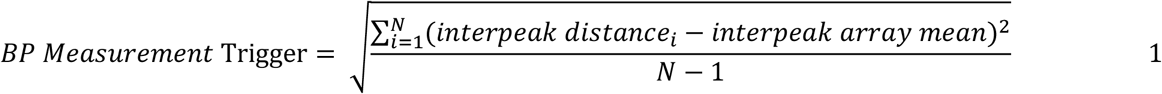

## Results and Discussion

The single participant clinical investigation was conducted according to the protocol and the results recorded and analysed.

### Results

The systolic, diastolic and pulse rate were measured with the Omron M3 Comfort according to the study protocol. The ECG and PPG data were analysed with the interpeak algorithm and new wait times calculated with code alterations as noted in Table 2; ^1^ standard deviation 10, ^2^ standard deviation 14.2, ^3^ interpeak minimum distance changed to 100 samples, ^4^ standard deviation 9.5 & ^5^ standard deviation 11.9

**Table 2:**
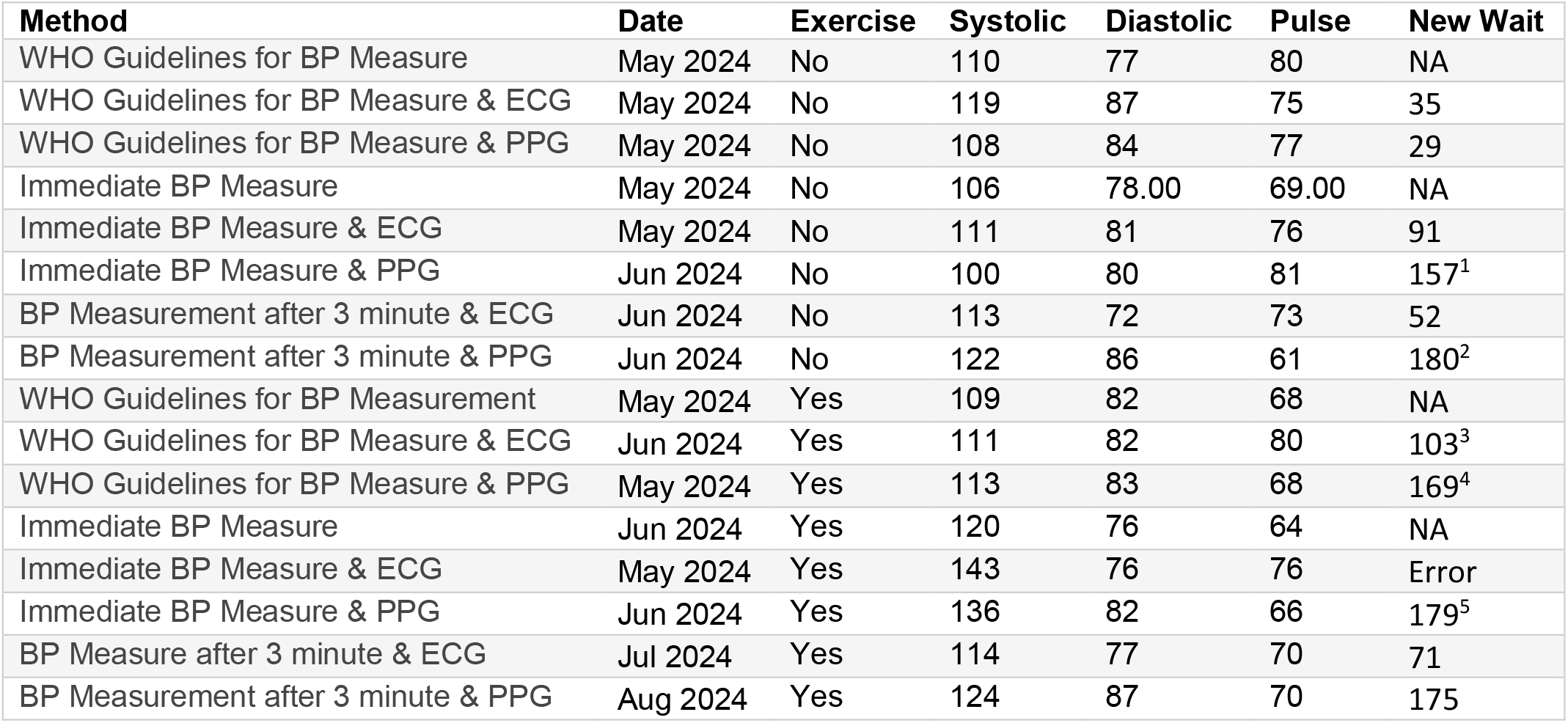
Single Participant Measurement Results.

| Method | Date | Exercise | Systolic | Diastolic | Pulse | New Wait |
| --- | --- | --- | --- | --- | --- | --- |
| WHO Guidelines for BP Measure | May 2024 | No | 110 | 77 | 80 | NA |
| WHO Guidelines for BP Measure & ECG | May 2024 | No | 119 | 87 | 75 | 35 |
| WHO Guidelines for BP Measure & PPG | May 2024 | No | 108 | 84 | 77 | 29 |
| Immediate BP Measure | May 2024 | No | 106 | 78.00 | 69.00 | NA |
| Immediate BP Measure & ECG | May 2024 | No | 111 | 81 | 76 | 91 |
| Immediate BP Measure & PPG | Jun 2024 | No | 100 | 80 | 81 | 157<sup>1</sup> |
| BP Measurement after 3 minute & ECG | Jun 2024 | No | 113 | 72 | 73 | 52 |
| BP Measurement after 3 minute & PPG | Jun 2024 | No | 122 | 86 | 61 | 180<sup>2</sup> |
| WHO Guidelines for BP Measurement | May 2024 | Yes | 109 | 82 | 68 | NA |
| WHO Guidelines for BP Measure & ECG | Jun 2024 | Yes | 111 | 82 | 80 | 103<sup>3</sup> |
| WHO Guidelines for BP Measure & PPG | May 2024 | Yes | 113 | 83 | 68 | 169<sup>4</sup> |
| Immediate BP Measure | Jun 2024 | Yes | 120 | 76 | 64 | NA |
| Immediate BP Measure & ECG | May 2024 | Yes | 143 | 76 | 76 | Error |
| Immediate BP Measure & PPG | Jun 2024 | Yes | 136 | 82 | 66 | 179<sup>5</sup> |
| BP Measure after 3 minute & ECG | Jul 2024 | Yes | 114 | 77 | 70 | 71 |
| BP Measurement after 3 minute & PPG | Aug 2024 | Yes | 124 | 87 | 70 | 175 |

The peak detect process was found to work optimally with the ECG sensor on the MAX86150EVSYS whilst the PPG sensor had significant bias and peak omission issues. A sample of ECG recordings taken whilst following the WHO guidelines is shown in Figure 2 below.

**Figure 2:**
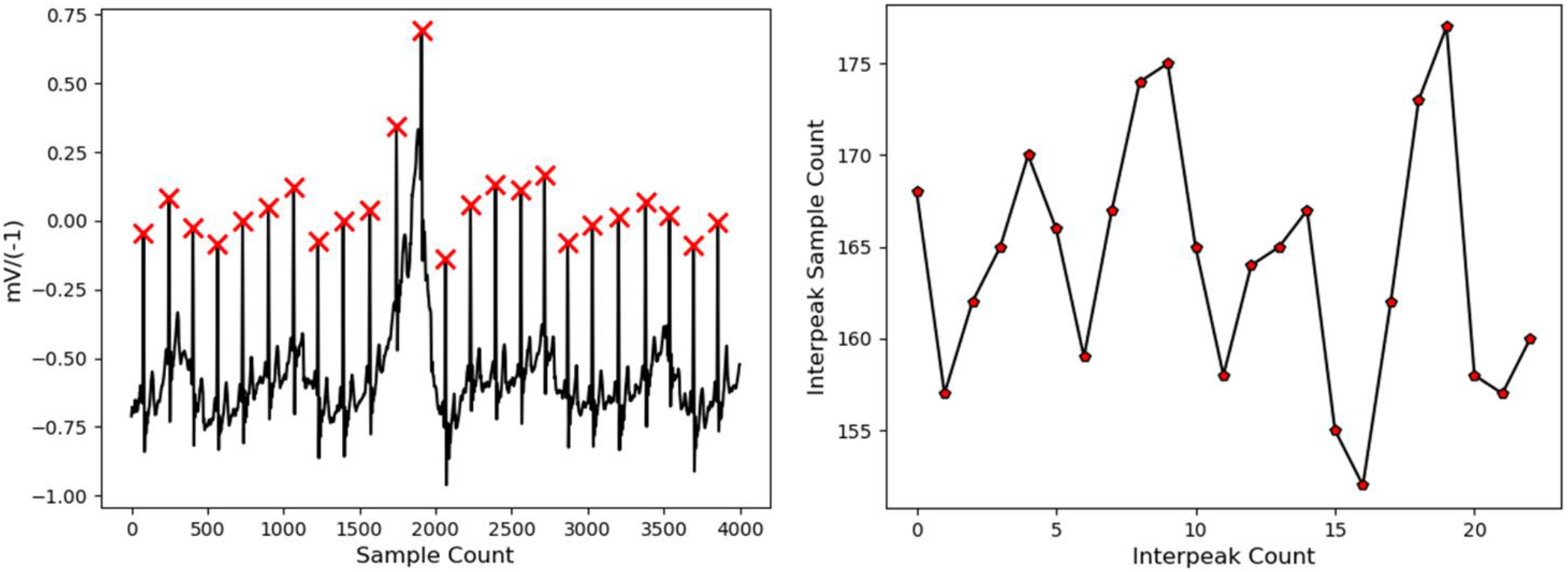
ECG data inverted on y axis with peak_detect(left) and interpeak array by sample count (right)

## Discussion

A heightened heart rate is associated with increased blood pressure (Scott & Drawz, 2012) and thus the exercise intervention was intended to increase BP into a region where measurement was unsuitable due the temporary nature of the elevated BP. The goal of the clinical investigation was to examine whether the point at which human BP returns to normal conditions could be determined using either photoplethysmography, (PPG) or electrocardiogram, (ECG), technology.

The WHO guideline compliant measurement with ECG monitoring and no intervention gave a new wait time of 35 seconds, representing an 88% reduction in wait time. The time to wait until the standard deviation conditions were met increased by 194% and 40% for the two ECG measurements in the case of intervention and reflected the expected trend. Due to a phenomenon known as heart rate variability, (HRV) (Tiwari, et al., 2022), the process of selecting a standard deviation threshold is non-trivial and further studies are required to validate the finding. Of note, is the corrective manner of HRV which seems to oscillate between extremes.

The resources for the clinical investigation were limited and several fields in which the study protocol could be improved were identified, namely personnel, research site, participants, intervention, duration, equipment communication and measurement method. In particular, the study participants should be expanded to include a more representative cohort of adults, with attention to age, gender, ethnicity, BMI and disease. In addition, the exercise intervention should be designed for a more sustained BP elevation and monitored to ensure precise pulse rate thresholds are achieved to ensure repeatability.

## Conclusions

The WHO guidelines specify a wait period of five minutes before taking a BP reading but literature suggests for healthy individuals that wait is unnecessary. Given the debate, lack of uniformity (Japan specify a three-minute wait) and the discretion that is ceded to healthcare professionals on their measurement approach, a more active and dynamic approach to correct BP measurement procedure is long overdue.

When the participant was waiting and complying with WHO guidelines, the clinical investigation using ECG data yielded a wait time reduction of 88%. Of note also, is the incremental corrective waveforms associated with heart rate variability and further study is recommended to determine where a correlation exists between HRV and trending blood pressure.

The single participant clinical investigation suggests that monitoring a patient’s ECG interpeak intervals could determine the most effective time to take a BP measurement. This procedure might successfully reduce the WHO mandated wait time of 5 minutes. A wait time of 35 seconds would likely be acceptable to clinicians and were the approach to be fully validated, it could represent a paradigm shift in healthcare where BP measurements become more reliable and repeatable and clinicians break the habit of disregarding good clinical practice guidelines.

This approach could be designed into standard oscillometric blood pressure cuff measurement devices with the addition of two electrodes on the outside of the device resulting in potentially significantly improved patient outcomes and increased blood pressure screening of patients.

## Data Availability

All data produced in the present study are available upon reasonable request to the authors

## Acknowledgements

I wish to acknowledge the support of the university faculty of TUS, in particular my supervisor Paul Morrow, course director Mairead Dennehy and dissertation lecturer Dr Lisa Henihan

## Notes

### Competing Interest Statement

The authors have declared no competing interest.

### Funding Statement

Funding received from Most Marvellous Company Ltd.

### Author Declarations

Ethics committee/IRB of Technological University of the Shannon gave ethical approval for this work

